# Co-Designing an Integrated Intervention Package to Strengthen Adolescent Sexual and Reproductive Health Services in Primary Healthcare Units in Southern Ethiopia: A Human-Centered Design Approach

**DOI:** 10.64898/2026.01.13.26344084

**Authors:** Negussie Boti Sidamo, Mengistu Meskele, Sultan Hussen Hebo, Abinet Teshome, Bilcha Oumer, Yenus Ibrahim, Menaye Yihune, Getnet Mitikie, Yalemzer Agegnehu

## Abstract

**Background:** Adolescent sexual and reproductive health (ASRH) remains a critical public health priority in Ethiopia, where adolescents face persistent barriers to accessing responsive and high-quality sexual and reproductive health (SRH) services. Conventional top-down interventions often fail to address adolescents’ lived experiences and local contexts. Human-centered design (HCD) offers a participatory and context-sensitive approach for developing adolescent-responsive interventions. As part of the **SAFE study** (***S***trengthening Prim***A***ry Healthcare ***F***aciliti***e***s), this study reports on the application of HCD to co-design an integrated ASRH intervention package for primary healthcare facilities in Southern Ethiopia.

**Methods:** The study was conducted from 25^th^ May to 5^th^ October 2025 in Arba Minch town and surrounding districts, encompassing urban and rural communities served by primary healthcare facilities. We employed a participatory co-design approach grounded in HCD and participatory action research to develop integrated intervention packages. This approach was selected to ensure contextual relevance, cultural sensitivity, and feasibility within existing health system structures. Adolescents, healthcare providers, youth leaders, facility managers, and researchers were engaged through iterative needs assessments, co-design workshops, and prototype refinement. Qualitative data were analyzed thematically and iteratively to inform Intervention package development.

**Results:** The co-design process generated four integrated intervention packages: (1) capacity building for healthcare providers and facility managers to enhance technical competence and adolescent-responsive service delivery; (2) peer navigator programs to enhance demand, service utilization, adherence, and linkage to ASRH service; (3) an SRH call center to Confidential information counseling, appointment, and bridge secondary barriers (barriers from treatment seeking to reach health service); and (4) strengthening adolescent-friendly health facilities to improve accessibility, privacy, and quality of care.

**Conclusion:** Human-centered co-design enabled the development of a comprehensive and integrated ASRH intervention package that addresses multilevel barriers Service provision, accessibility, utilization, and integration. Institutionalizing human-centered approaches within existing health systems may enhance equity, effectiveness, and sustainability of ASRH services. Future research should evaluate the implementation, scalability, and impact of these interventions.

## Introduction

Adolescence is a pivotal stage of development characterized by rapid physical, psychological, and social transitions that shape lifelong health and well-being(1). While the WHO defines adolescence as ages 10–19 years, recent evidence indicates that extending this range to 10–24 years more accurately reflects biological and social development and enables interventions to reach a broader group of young people, particularly in improving access to sexual and reproductive health services (1-3). Expanding the age range acknowledges the extended period of vulnerability and opportunity for intervention, especially in low-resource settings.

Despite being a critical period for establishing lifelong health behaviors, yet young people in low- and middle-income countries face persistent barriers to accessing sexual and reproductive health (SRH) services(4). Key challenges include limited availability of adolescent-responsive care, social stigma, lack of privacy, and fragmented service delivery (5-7). In Ethiopia, specifically, where adolescents constitute over one-fifth of the population, these barriers are particularly pronounced, encompassing provider bias, insufficient privacy and confidentiality, limited access to contraceptives, and pervasive stigma surrounding adolescent sexuality(9-11). As a result, these factors constrain both utilization and effectiveness of adolescent SRH services, leaving young people at heightened risk of early and unintended pregnancies, unsafe abortions, sexually transmitted infections (STIs), and other adverse health and social outcomes(14).

Conventional top-down SRH interventions have repeatedly failed to address adolescents’ lived experiences, largely because young people are seldom involved in the design or implementation of health services(1). This exclusion often results in interventions that are poorly aligned with adolescents’ needs, preferences, and sociocultural realities, leading to limited acceptability and suboptimal utilization(2). In Ethiopia, existing adolescent SRH programs and studies have predominantly adopted expert-driven or provider-centered approaches, with minimal use of adolescent participatory design methods(3). Moreover, much of the available evidence focuses on intermediate outcomes such as knowledge, attitudes, or service uptake, while paying limited attention to implementation dimensions including feasibility, scalability, fidelity, and sustainability within primary healthcare systems (4, 5). Together, these patterns illustrate the persistent shortcomings of top-down approaches and the resulting disconnect between program design and adolescents’ everyday realities.

To address the limitations of conventional top-down approaches, innovative and context-sensitive strategies are needed that actively engage adolescents and other stakeholders in the design and implementation of SRH interventions (1). Human-Centered Design has emerged as a promising alternative, offering a structured yet flexible framework for co-creating interventions that are responsive to adolescents’ needs, preferences, and lived experiences (2-4). By centering the voices and experiences of adolescents, healthcare providers, youth leaders, and facility managers, HCD-informed interventions can enhance relevance, acceptability, feasibility, and effectiveness, while explicitly addressing both supply- and demand-side barriers. Unlike conventional top-down models, HCD fosters solutions that are more likely to be feasible, scalable, and sustainable within local health system constraints(5). Integrating HCD into adolescent SRH programming therefore holds significant potential to improve equity, utilization, and health outcomes, particularly in resource-constrained settings such as Ethiopia. Despite this promise, few studies in Ethiopia have systematically applied participatory or human-centered approaches that meaningfully involve adolescents and community stakeholders, highlighting a critical gap in both research and practice (4, 6).

Given this evidence gap, there remains limited understanding of how to effectively design and implement adolescent-responsive SRH interventions in primary healthcare settings in Ethiopia. In response, the present study employed a co-design process to develop an integrated package of interventions aimed at strengthening the accessibility, quality, and acceptability of SRH services for adolescents. By actively engaging adolescents, healthcare providers, and facility managers in the design process, this study sought to ensure that the resulting interventions are contextually relevant, feasible, and responsive to the unique needs of Adolescents in Southern Ethiopia.

## Methods

### Study Setting

This study was conducted in Arba Minch and surrounding districts, covering both urban and rural communities served by primary healthcare facilities. These facilities provide essential SRH services but face persistent challenges, including low service utilization, limited adolescent and youth-friendly spaces, gaps in provider training, and resource constraints. Service delivery is further complicated by inadequate privacy, periodic stock-outs of essential commodities, and prevailing sociocultural norms that influence adolescents’ health-seeking behaviors. Understanding these multifaceted contextual realities is critical to designing interventions that are responsive, acceptable, and effective for adolescents in these settings.

### Study Design and Period

This study employed a participatory co-design approach, grounded in the principles of human-centered design (HCD) and participatory action research, to develop intervention packages tailored to the needs of adolescents and young people. Co-design was deliberately chosen to ensure interventions were locally relevant, culturally sensitive, and feasible within existing health system structures. The research followed iterative learning cycles, with the Design Phase utilizing HCD methods to anchor interventions in user experiences, technical feasibility, and alignment with routine service delivery. Central to this phase were three participatory workshops encompassing all five stages of the HCD process. These workshops facilitated active stakeholder engagement, fostered collaborative problem-solving, and enabled the co-creation of practical, scalable, and context-driven solutions within primary healthcare settings. Participant recruitment started on 25/05/2025 and ended on 05/10/2025.

### Participants and Stakeholders

The co-design process engaged a diverse, purposively selected group of stakeholders, including adolescents (10 to 24 years), youth leaders, healthcare providers, facility managers, senior researchers, and the design research team. Selection prioritized representation across age, gender, professional roles, and decision-making levels relevant to ASRH service delivery. Adolescents and community youth leaders contributed experiential insights on barriers to access and preferences for care, while healthcare providers and facility managers offered perspectives on service delivery constraints, workforce capacity, and implementation feasibility. Senior researchers and the design team provided Prototype development throughout the co-design activities. Purposive and snowball sampling ensured a wide range of voices across geographic and social contexts, enabling a rich, multi-level understanding of challenges and opportunities for strengthening ASRH services.

### Human-Centered Design Framework

The development of interventions was guided by a **HCD framework**, implemented through three iterative and complementary phases. **Phase 1** involved a deep exploration of adolescents’ experiences and perspectives to uncover critical barriers to accessing sexual and reproductive health (SRH) services. **Phase 2** translated these insights into “How Might We” (HMW) questions, stimulating creative, context-driven solution generation. Emerging solutions were then thematically organized and rigorously assessed for feasibility, acceptability, and potential impact. **Phase 3** emphasized prototyping and iterative testing with key stakeholders including adolescents, healthcare providers, and facility managers—allowing interventions to be refined based on real-world feedback. This approach ensured that the final interventions were practical, culturally relevant, and fully aligned with the capacities and realities of the local health system.

## Co-Design Activities

The co-design process was conducted through a series of iterative, structured stages guided by HCD principles. Prior to initiating the workshops, findings from previous studies in the study area were reviewed to inform the design and focus of the co-design activities. Building on this evidence, three participatory HCD workshops were conducted with adolescents, healthcare providers, and facility managers.

The first workshop (Co-Discovery/Empathy) aimed to explore barriers to adolescent SRH service access and utilization. Participants collaboratively mapped challenges through participatory discussions and reflections. In parallel, in-depth interviews (IDIs) with healthcare providers and facility managers examined service delivery constraints and contextual factors influencing adolescent care. Facility assessments documented infrastructure, resources, and service delivery practices relevant to adolescent SRH. Collectively, these activities generated a comprehensive understanding of barriers and opportunities for strengthening adolescent-friendly SRH services.

The second workshop (Co-Design) focused on solution generation. Using “How Might We” (HMW) questions, participants—including adolescents, youth representatives, healthcare providers, facility managers, and academic researchers engaged in brainstorming, journey mapping, and role-play exercises to foster creativity and collective problem-solving. Proposed solutions were then clustered thematically and prioritized according to feasibility, acceptability, and alignment with existing health system Using Google docs. The third workshop (Prototyping) translated selected ideas into low-fidelity prototypes, such as Call center, peer navigator initiatives, adolescent-friendly service modifications, community engagement strategies, and health learning materials. Prototypes were iteratively refined through structured feedback loops with participants to ensure practicality, contextual relevance, and alignment with service delivery realities. The fourth stage (Pilot Testing) involved implementing the prototypes on a small scale to evaluate feasibility, acceptability, and appropriateness within routine primary healthcare settings.

The fifth stage (Iterative Refinement and Integration) consolidated lessons learned from pilot testing into a cohesive, integrated intervention package, which was subsequently validated through consultative meetings with adolescents, facility managers, and district health officials. The final package was aligned with national adolescent and youth health guidelines to ensure policy relevance and sustainability. All workshops and interviews were conducted in the local language by trained qualitative researchers. Sessions were audio-recorded with participants’ consent, transcribed verbatim, and supplemented with detailed field notes to capture non-verbal cues and contextual nuances. Data sources included facilitator notes, flip charts, visual artifacts, and workshop outputs, ensuring that the co-design process was grounded in a rich and nuanced understanding of adolescents’ experiences and service delivery realities.

### Data Analysis

Data from the co-design process were analyzed using a systematic qualitative synthesis approach. Generated ideas from workshops were first organized through clustering and affinity mapping, followed by thematic analysis to identify recurring patterns and intervention domains. Qualitative data from in-depth interviews, workshops, and field observations were transcribed verbatim, translated into English when necessary, and coded inductively. The coding framework was iteratively refined to capture barriers, facilitators, and opportunities for strengthening ASRH services.

Proposed solutions were then clustered thematically and prioritized according to feasibility, acceptability, and alignment with existing health system structures. Disagreements were resolved through facilitated group discussions and consensus-building exercises, ensuring that the final intervention packages reflected shared priorities across stakeholder groups. Workshop outputs were examined through both socio-ecological and health systems lenses, allowing analysis across individual, facility, community, and policy levels. Findings were organized into thematic categories and further prioritized according to feasibility, potential impact, urgency, stakeholder support, and alignment with study objectives. Participatory HCD tools, including storytelling, journey mapping, and feasibility–impact matrices, were employed to validate and refine key insights. Feedback from prototypes and co-designed solutions was triangulated with interview and facility assessment data, capturing both convergent and divergent perspectives. Discrepancies were resolved through consensus discussions, while field notes and reflective memos enriched the contextual interpretation of findings.

## Results

### Participant Characteristics

A total of 63 participants were engaged in the co-design process, representing diverse stakeholder groups. Among these, 35 were adolescents and young people aged 10–24 years, with an approximately equal distribution of males and females, recruited from rural, urban and semi-urban communities. Adolescents were purposively selected to capture variation in school attendance, residence in hotspot areas, and prior experience with health services. In addition, the study included eight healthcare providers (nurses, midwives, and health extension workers) and four facility managers, who contributed insights into service delivery practices and health system challenges. The process also involved eight research team members, four senior researchers, and four local health officials, who participated in consultative workshops to ensure contextual relevance and alignment with local priorities and policy frameworks. Overall, the sample provided a balanced mix of end-users, service providers, and stakeholders, supporting a comprehensive understanding of barriers, facilitators, and opportunities for strengthening ASRH services in primary healthcare settings.

### Key Findings from the Co-Design Process

The co-design workshops and complementary activities generated rich insights into the barriers, facilitators, and opportunities for improving adolescent SRH services. Findings are presented thematically, reflecting both the stages of the HCD process,, socio-ecological model, and health systems levels of influence.

#### Barriers to Adolescent SRH Service Utilization

The design team initially convened virtually via Zoom, led by the project lead, to review formative research findings and collaboratively identify barriers to ASRH service access and utilization. This was followed by a face-to-face *Empathize & Define Problem Workshop*, which included adolescents, healthcare providers, facility managers, and design team members from both rural and semi-urban communities.

The workshop began with a presentation of formative research findings, synthesizing perspectives from Adolescents and young people, service providers, facility managers, senior researchers, and design team members regarding SRH needs, barriers, and existing services. Analysis employed a hybrid framework combining the SEM with a health systems lens, providing a comprehensive, multi-dimensional understanding of factors shaping ASRH Accessibility, utilization, and adherence. The SEM highlighted barriers across five interrelated levels intrapersonal, interpersonal, institutional, community, and structural capturing the complex interplay of personal, social, and environmental influences on Adolescent service seeking, utilization and compliance behavior. Complementing this, the health systems perspective identified operational and systemic challenges, including provider capacity, facility readiness, accessibility, and governance limitations. This dual approach ensured findings were both contextually grounded, reflecting adolescents’ lived experiences, and structurally informed, addressing broader health system constraints affecting service delivery.

Participants initially identified 57 barriers, which were subsequently grouped into three interrelated levels: individual/intrapersonal, family/community, and health system/structural. A facilitated discussion then validated the findings, ensuring they accurately reflected stakeholders’ experiences and surfaced overlooked perspectives. Through an interactive prioritization exercise, 19 key barriers were selected for ideation workshop. **(Appendix table 1)**.

### Ideation Workshop Findings

The Ideation Workshop, the second phase of the process, focused on translating the prioritized ASRH barriers identified in Phase 1 into actionable solutions. The workshop convened a diverse group of stakeholders including adolescents and youths, healthcare providers, youth leaders, researchers, and the design team who collaboratively refined and finalized priority intervention areas. From the 19 barriers initially identified through expert consultation, 10 priority barriers to ASRH service access and utilization were validated based on severity, feasibility of intervention, and consistency with the lived experiences of adolescents and service providers. These barriers were clustered into three thematic areas: Service Delivery; Family and Community Engagement; and Information and Education. This thematic grouping provided a structured framework for intervention design.

Using the **“How Might We…” (HMW) framework**, participants collectively reframed these thematic barriers into solution-oriented problem statements. This process merged overlapping challenges, addressed underlying root causes, and transformed problem-centered discussions into opportunity-driven exploration. The resulting HMW questions guided the development of culturally appropriate, contextually relevant, and adolescent-responsive interventions across the three thematic areas. Thus, this streamlined ideation process ensured that subsequent intervention design was evidence-informed, stakeholder-driven, and user-centered, establishing a strong foundation for prototyping and testing contextually appropriate ASRH solutions**(Appendix table 2)**.

## Idea Generation and Thematic Clustering

During the co-design process, participants generated a total of 140 creative and contextually grounded ideas in response to the priority barriers affecting access to and utilization of ASRH services. The ideation session, guided by the previously developed ***“How Might We…questions”*** fostered divergent thinking and drew on the diverse perspectives of adolescents and young people, youth leaders, service providers, facility managers, senior researchers, and members of the design team. The ideas multispectral network digital innovations, peer-to-peer support models, health system strengthening strategies, and community-based interventions, reflecting a comprehensive and inclusive approach to addressing ASRH barriers.

Following generation, participants engaged in a collaborative clustering exercise to organize the ideas into three strategic thematic areas. **Service Delivery** (71 ideas) focused on improving access, promoting youth-friendly services, enhancing provider capacity, and strengthening overall service quality. **Family and Community Engagement** (40 ideas) emphasized parental involvement, fostering supportive community environments, and ensuring culturally responsive interventions. **Information and Education** (29 ideas) highlighted strategies to improve adolescents’ knowledge, increase awareness, and optimize communication channels. The clustering process was guided by explicit criteria, including alignment with objectives and intended outcomes, relevance to target populations, feasibility given existing resources, cultural appropriateness, potential impact on ASRH outcomes, and accountability of implementing actors. Each cluster was reviewed and presented to the plenary, with clear rationales linking the grouped ideas to the identified ASRH barriers. This systematic approach enabled participants to synthesize contributions, identify emerging solution patterns, eliminate redundancies, and distill actionable strategies. Overall, the session produced dozens of innovative, contextually grounded ideas, organized into coherent thematic clusters, providing a strong foundation for the design of integrated interventions in the next phase of the project. **(Appendix table 3)**.

### Feasibility and Impact Assessment

Feasibility-Impact Matrix was used to prioritize intervention themes based on technical feasibility, resource requirements, potential impact, and scalability. **Service Delivery Strengthening** emerged as both highly feasible and high impact, with key strengths including being actionable, infrastructure-ready, and scalable; implementation challenges primarily involved Monitoring and evaluation training, making it a top priority for immediate implementation (Phase 1). **Information, Education, and Communication (IEC) Strategies** were rated moderate-to-high for feasibility and impact, with the advantage of being scalable and adaptable to digital or peer-led platforms; however, potential barriers such as digital access disparities and literacy gaps suggested their phased implementation between Phase 1 and Phase 2. **Family and Community Engagement** was considered moderate in feasibility but high in impact, as it fosters supportive norms and encourages community ownership, though it requires sustained behavioral change and is therefore suitable for later phases of implementation (Phase 2 onward). Overall, this assessment provided a structured approach to balancing practicality and potential impact, guiding the strategic rollout of adolescent sexual and reproductive health interventions. **(Appendix table 4)**.

### Developing final intervention package

Following the Feasibility and Impact Assessment, prioritized themes were translated into actionable strategies through a co-design process. The process began with mapping specific activities to the high-priority areas of service delivery strengthening, information and education strategies, and family and community engagement, ensuring contextual relevance and stakeholder acceptability. Resource requirements were assessed, and a phased implementation plan aligned high-feasibility interventions with immediate rollout and moderate-feasibility interventions with later phases, supported by a monitoring and evaluation framework to guide iterative refinement.

A Prototype Workshop further refined the interventions by translating prioritized solutions into low-fidelity prototypes to assess usability, acceptability, and alignment with adolescent needs. This included role-play simulations of adolescent-provider interactions, visual mock-ups of youth-friendly spaces and educational materials, and service flow diagrams of integrated SRH pathways. From this process, five key prototypes were selected for pilot implementation: capacity-building for providers and managers, an integrated peer navigation program, SRH call center support, adolescent-friendly service environments, and a supportive supervision and mentorship system. This stepwise, co-designed approach ensures that interventions are contextually grounded, strategically phased, and provide a comprehensive, multi-level framework to strengthen adolescent sexual and reproductive health services.

### Description of Final Intervention Packages

Through an iterative co-design and prototyping process, four integrated intervention packages were developed to strengthen ASRH services in primary healthcare facilities. Each package addresses distinct but complementary barriers at the individual, provider, facility, and community levels, incorporating stakeholder input to ensure contextual relevance, feasibility, and sustainability. The first package, **Capacity-Building for Providers and Managers**, includes in-person workshops, interactive sessions, and on-site mentorship to equip health providers and facility managers with competencies in integrated and adolescent-friendly SRH service delivery. The training focuses on VCAT, non-judgmental communication, confidentiality, and client-centered care, with these principles reinforced through continuous supervision, periodic refresher sessions, and structured performance feedback. The second package, **Adolescent-Friendly Service Environments**, focuses on operational modifications within facilities to improve comfort, privacy, and accessibility for adolescents. Adjustments include welcoming waiting and consultation areas, extended service hours to accommodate school schedules, adolescent-specific service blocks to reduce waiting times, and visual Health learning materials such as banners and leaflets that promote inclusivity, confidentiality, and awareness of available services. The third package, **Integrated Peer Navigation** targets hotspot areas by engaging peer navigators to provide SRH information, counseling, and guidance through outreach activities, facility-based support, and community follow-ups. Structured referral pathways link adolescents to appropriate health facilities and social services, while continuous monitoring ensures effective engagement and utilization of services, fostering a supportive peer-driven network. The fourth package, **SRH Call Center Support**, establishes a confidential toll-free hotline to provide real-time SRH information, counseling, appointment scheduling, and referral linkages. Call center staff are trained in adolescent communication, privacy protocols, and accurate information dissemination, with integration into facility records ensuring follow-up and continuity of care. During implementation, the call center received numerous inquiries primarily regarding contraception, menstrual health, sexually transmitted infections, and confidentiality concerns. Adolescents reported high satisfaction with the service, citing convenience, anonymity, and reliability, while facility records indicated increased in-person service utilization. **(Appendix table 5)**.

## Discussion

This study co-designed an integrated intervention package to strengthen ASRH services using a human-centered design approach. Four complementary interventions provider capacity-building, peer navigation, an SRH call center, and adolescent-friendly health facilities were developed to address multilevel barriers to ASRH service utilization, spanning system-, community-, and individual-level challenges identified through the human-centered design process. Engaging adolescents, service providers, and community stakeholders throughout the co-design process ensured that the interventions were contextually appropriate, acceptable, and feasible. The iterative co-design and prototyping approach enhanced both the relevance and practical applicability of these interventions while fostering stakeholder ownership and alignment with local needs, health system structures, and national guidelines. These findings demonstrate that participatory, human-centered design approaches can produce integrated, tailored interventions capable of addressing complex barriers to ASRH service access and utilization.

The capacity-building package for healthcare providers and facility managers emphasized VCAT, aiming to strengthen technical competence, enhance communication skills, and foster adolescent-responsive attitudes. Consistent with prior research, inadequate provider capacity and judgmental behaviors remain major barriers to ASRH service utilization; systematic reviews and country-level studies have shown that negative provider attitudes and limited competency discourage adolescents from seeking care and undermine youth-friendly service delivery(21-23).

By addressing these challenges, the package enhances service quality and fosters an enabling environment for adolescents, which is critical for sustaining service uptake(19). These findings underscore the importance of ongoing capacity-building efforts, guided by the identification of existing gaps, to maintain and further improve adolescent-responsive care.

The integration of SRH call center and peer navigator packages was designed to reach hard-to-access adolescents, providing convenient, timely information and support while fostering strong community and facility networks. The peer navigator approach leveraged adolescents’ preference for guidance from trusted peers, serving as a bridge between adolescents and health facilities to overcome social and informational barriers, consistent with evidence that peer-led interventions improve ASRH knowledge, reduce stigma, and facilitate timely service linkage. Complementing this, the SRH call center provided confidential, real-time sexual and reproductive health information and counseling, offering anonymity and remote access that are particularly valuable in contexts where stigma and fear of disclosure limit service utilization. These findings are supported by prior studies: in Zambia, community-based, peer-led SRH services significantly increased adolescents’ knowledge of their HIV status and access to SRH services (24). while pilot studies in Kenya demonstrated that peer navigators engaging marginalized youth led to high acceptance of HIV testing and improved linkage to treatment when peers accompanied adolescents through the care process (25). Moreover, reviews of mHealth and digital interventions indicate that mobile and Internet-based approaches effectively improve adolescents’ SRH knowledge and provide confidential, tailored information beyond traditional settings (26). These findings imply that combining peer support with digital platforms can effectively enhance adolescents’ access to accurate SRH information and timely services.

Strengthening adolescent-friendly health facilities was central to ensuring that increased demand translated into effective service utilization. As part of this package, we designed dedicated adolescent waiting areas and welcoming rooms, established adolescent clinics, provided informational brochures in local languages, ensured privacy, promoted respectful care, and offered convenient service hours. Aligning services with these adolescent-friendly standards has been consistently associated with higher satisfaction and greater service uptake(14). Another systematic reviews in Ethiopia show that facility features such as convenient working hours and proximity are strongly associated with increased utilization of SRH services among youth(27). These findings imply that by reinforcing the supply-side readiness of the health system, such interventions create an environment conducive to sustained adolescent engagement with sexual and reproductive health services.

Using Human-Centered Design (HCD) to actively engage adolescents, youth leaders, healthcare providers, facility managers, and local authorities throughout the co-design process ensured that interventions effectively addressed real-world barriers. Iterative feedback during prototyping and pilot testing further refined the interventions, enhancing alignment with facility workflows and community norms. These results are consistent with previous studies conducted in diverse settings, highlighting the benefits of active stakeholder participation in fostering ownership and motivation, which in turn increases the likelihood of sustained implementation beyond the study period (17, 19, 28). Consistent with findings from the Adolescents 360 project across three successful countries, this demonstrates HCD’s value in designing context-responsive interventions that promote ownership, sustainability, and scalability (18, 29). Similarly, evidence from Kenya shows that applying HCD processes when translating evidence-based interventions builds on valuable knowledge while ensuring the intervention meets the needs of the new target audience(30). Effective adolescent SRH programming, therefore, requires multi-level strategies that address individual behaviors, community norms, and health system capacities, with phased implementation optimizing resources and achieving sustainable gains (19, 31). Collectively, these findings underscore that co-designed, system-level interventions can meaningfully improve adolescent SRH outcomes and inform scalable strategies in similar low-resource settings (32). They also have important implications for policy and program development: aligning interventions with national adolescent health guidelines facilitates scale-up and integration into routine primary healthcare services, while participatory co-design approaches promote stakeholder ownership and sustainability, offering a practical framework that maximizes feasibility, acceptability, and contextual fit—key considerations for future program planning and evaluation (8, 33)

### Strengths and Limitations

This study has several key strengths. The use of participatory co-design and human-centered design approaches, combined with multi-stakeholder engagement, iterative prototyping, and triangulation of multiple data sources, enhanced the validity, relevance, acceptability, and feasibility of the interventions. Engaging adolescents, healthcare providers, facility managers, and community stakeholders throughout the design process ensured that the intervention packages reflected users’ lived experiences, preferences, and constraints an approach increasingly recognized as best practice in adolescent health programming. Notably, the four intervention packages functioned synergistically: capacity building and adolescent-friendly facilities strengthened the supply side, while peer navigators and the SRH call center addressed demand generation and access barriers. By integrating community-based, digital, and facility-level interventions, the study employed a comprehensive health systems approach that is more likely to achieve sustainable improvements in ASRH service delivery. These findings imply that co-design and human-centered approaches enhance the contextual relevance, acceptability, and feasibility of adolescent health interventions, offering a scalable model with potential applicability to other low-resource settings. However, Limitations include the potential limited generalizability of findings beyond the selected facilities in Southern Ethiopia and the fact that pilot testing was conducted in only a subset of settings, which may not capture all operational challenges in diverse contexts.

## Conclusion

Integrating co-design and HCD approaches into ASRH interventions substantially enhances contextual relevance, acceptability, and feasibility, while effectively addressing multi-level barriers to service utilization. This study demonstrates that HCD is a powerful strategy for strengthening adolescent SRH services in Ethiopia by actively engaging adolescents, providers, and facility managers in identifying challenges and co-creating solutions. The process produced four practical, low-cost prototypes provider capacity building, peer navigators, a confidential call center, and adolescent-friendly service environments that collectively offer a scalable and sustainable model for improving adolescent health services in low-resource settings. By combining structural enhancements with demand-side strategies, these interventions extend service reach beyond facility walls, mitigate stigma, improve accessibility, and provide timely, confidential support. Unlike conventional interventions targeting isolated components, this integrated, system-level approach illustrates how co-designed, contextually adapted packages can achieve broader coverage, stronger engagement, and measurable impact, providing a practical blueprint for national scale-up. Future research should prioritize piloting and rigorous evaluation of these prototypes to assess effectiveness, cost-efficiency, and long-term sustainability. Overall, HCD holds transformative potential to advance adolescent SRH service delivery and to accelerate progress toward national and global adolescent health goals.

### Policy and Practical Implications

This study provides compelling evidence that co-designed; HCD interventions can overcome multi-level barriers to ASRH service utilization, enhancing accessibility, acceptability, and engagement. **Policy implications** are clear: national ASRH strategies should formally integrate HCD and participatory co-design principles to ensure interventions are contextually relevant, adolescent-centered, and responsive to real-world challenges. Policymakers should prioritize system-level, multi-component approaches that combine structural improvements such as adolescent-friendly facilities and provider capacity building with demand-side strategies like peer navigation and confidential call centers, thereby maximizing reach and sustainability.

**For program implementers**, the findings underscore the importance of actively engaging adolescents, healthcare providers, facility managers, youth leaders and researchers throughout intervention design and implementation. Iterative prototyping and pilot testing enable alignment with facility workflows and community norms, strengthening feasibility, ownership, and long-term uptake. The low-cost, scalable prototypes developed—provider capacity building, peer navigators, SRH call centers, and adolescent-friendly service environments—offer a practical blueprint for replication in low-resource settings.

**Scalability and sustainability** can be achieved by integrating these interventions into routine primary healthcare services, phasing implementation to optimize resources, and evaluating cost-effectiveness and long-term impact. Collectively, these findings provide actionable guidance for policymakers, program implementers, and researchers seeking to design and scale effective, adolescent-centered SRH services.

#### Ethical Considerations

This study was conducted in accordance with the Declaration of Helsinki. Ethical approval was obtained from the Institutional Review Board (IRB) of Arba Minch University, College of Medicine and Health Sciences (Approval No: IRB/23339/25) on May 20^th^, 2025, prior to data collection. Written informed consent was obtained from all participants. For participants under 18 years, consent was obtained from parents or legal guardians, and assent was obtained from the adolescents. Participants aged 18 years and above provided their own written consent.

To ensure confidentiality, privacy, and anonymity, identifying information was removed from all reports, and participant identifiers were used only by the research team for internal purposes. Data collection occurred in private settings, participation was voluntary, and participants were informed of their right to withdraw at any time without consequences. All reported data were anonymized to prevent identification by family members, community members, or healthcare providers.

## Acknowledgments

The authors gratefully acknowledge the financial support provided by the IPHC-E Small Grants initiative, which made this study possible. We also thank Arba Minch University for providing the IRB approval letter. Our sincere appreciation goes to all study participants and data collectors for their valuable time, insights, and contributions. Additionally, the authors acknowledge the use of **ChatGPT-4** for language editing, organization of qualitative themes, and formatting. All AI-assisted outputs were rigorously reviewed and verified by the authors to ensure accuracy, clarity, and scholarly integrity.

## Data Availability

All data underlying the findings of this study are fully available without restriction and are included within the manuscript and its Supporting Information files. Additional data are available from the corresponding author upon reasonable request.

## Authors’ Contributions

All authors contributed equally to all stages of this work, including study conceptualization and design, data collection and analysis, interpretation of findings, and manuscript drafting. All authors reviewed, edited, and approved the final version of the manuscript.

## Conflict of Interest

All authors declare that they have no conflicts of interest.

Table captions

**Appendix Table 1:** Key Barriers to Adolescent Sexual and Reproductive Health Service Utilization

**Appendix Table 2:** Priority Adolescent Sexual and Reproductive Health Problems by Theme

**Appendix Table 3:** Participant-Generated Ideas and Thematic Clusters

**Appendix Table 4:** Feasibility and Impact Assessment of Adolescent Sexual and Reproductive Health Intervention Themes

**Appendix Table 5:** Final Co-Designed Adolescent Sexual and Reproductive Health Intervention Packages Designed in South Ethiopia, 2025.

## References

1. Sawyer SM, Azzopardi PS, Wickremarathne D, Patton GC. The age of adolescence. The lancet child & adolescent health. 2018;2(3):223–8.

2. Kinghorn A, Shanaube K, Toska E, Cluver L, Bekker L-G. Defining adolescence: priorities from a global health perspective. The Lancet Child & Adolescent Health. 2018;2(5):e10.

3. Guthold R, Stevens GA, Riley LM, Bull FC. Global trends in insufficient physical activity among adolescents: a pooled analysis of 298 population-based surveys with 1· 6 million participants. The lancet child & adolescent health. 2020;4(1):23–35.

4. Erasmus MO, Knight L, Dutton J. Barriers to accessing maternal health care amongst pregnant adolescents in South Africa: a qualitative study. International journal of public health. 2020;65(4):469–76.

5. Alkhidir ENM, Nouraldein E, Doran R. Challenges, barriers and facilitators contributing to progress towards achieving SDG 3.7: a documentary review of selected Sub-Saharan African countries: Thammasat University; 2024.

6. Ghimire R, Sharma S, Singh A, Bhusal R, Tamang L. Barriers to accessibility and availability of safe abortion services among young women in Nepal: a mixed-methods study. BMC Public Health. 2025;25(1):4113.

7. Ninsiima LR, Chiumia IK, Ndejjo R. Factors influencing access to and utilisation of youth-friendly sexual and reproductive health services in sub-Saharan Africa: a systematic review. Reproductive health. 2021;18(1):135.

8. Vandermorris A, Wigle J, Tam M, Peresin J, Dalal S, Kwong I, et al. Application of Youth-Led Participatory Action Research to Examining Adolescent Sexual and Reproductive Health and Rights in Ontario: What Can We Learn? Health Promotion Practice. 2025;26(5):913–25.

9. Seid A, Yeneneh H, Sende B, Belete S, Eshete H, Fantahun M, et al. Barriers to access safe abortion services in East Shoa and Arsi Zones of Oromia Regional State, Ethiopia. The Ethiopian Journal of Health Development. 2015;29(1).

10. Sidamo NB, Kerbo AA, Gidebo KD, Wado YD. Exploring barriers to accessing adolescents sexual and reproductive health Services in South Ethiopia Regional State: a phenomenological study using Levesque’s framework. Adolescent Health, Medicine and Therapeutics. 2024:45–61.

11. Wakjira DB, Habedi D. Barriers to access and utilisation of sexual and reproductive health services among adolescents in Ethiopia: a sequential mixed-methods study. BMJ open. 2022;12(11):e063294.

12. Zolfaghari E, Armaghanian N, Waller D, Medlow S, Hobbs A, Perry L, et al. Implementation science in adolescent healthcare research: an integrative review. BMC Health Serv Res. 2022;22(1):598.

13. Feyisa KW, Bayou NB, Siraneh Y, Feyisa BW. Process evaluation of the adolescent and youth health program implementation in primary healthcare facilities of Woliso town, Oromia regional state, Ethiopia: a case study evaluation design. Reproductive Health. 2025;22(1):196.

14. Health FDRoEMo. National Adolescents and Youth Health Strategy (2021–2025). Federal Democratic Republic of Ethiopia Ministry of Health; 2021.

15. Organization WH. WHO recommendations on adolescent sexual and reproductive health and rights. 2018.

16. Lassi ZS, Neideck EG, Aylward BM, Andraweera PH, Meherali S. Participatory action research for adolescent sexual and reproductive health: a scoping review. Sexes. 2022;3(1):189–208.

17. Wallace LJ, Darko NA, Diarra A, Yaogo M, Gyawu B, Prempeh P, et al. Co-creating gender-transformative interventions for adolescent mental, sexual, and reproductive health and rights. African Journal of Reproductive Health/La Revue Africaine de la Santé Reproductive. 2025;29(6s):20–35.

18. Wilson M, Cutherell M, Musau A, Malakoff S, Coppola A, Ayenekulu M, et al. Implementing adaptive youth-centered adolescent sexual reproductive health programming: learning from the Adolescents 360 project in Tanzania, Ethiopia, and Nigeria (2016-2020). Gates Open Research. 2022;6:14.

19. Cutherell M, Coppola A, Ogbondeminu F, Syomiti B, Atlie S, Mtei E, et al. Data-driven program adaptation and continuous improvement: A case study of the Adolescents 360 (A360) project in Ethiopia, Nigeria, and Tanzania. VeriXiv. 2025;1:2.

20. Fasil N, Worku A, Oljira L, Tadesse AW, Berhane Y. Effect of peer-group participation on Knowledge about condoms among adolescent girls in rural Eastern Ethiopia: a community-based repeated cross-sectional study. medRxiv. 2024:2024.12. 12.24318896.

21. Onukwugha FI, Hayter M, Magadi MA. Views of Service Providers and Adolescents on Use of Sexual and Reproductive Health Services by Adolescents: A Systematic Review. African journal of reproductive health. 2019;23(2):134–47.

22. Sidamo NB, Kerbo AA, Gidebo KD, Wado YD. Socio-ecological analysis of barriers to access and utilization of adolescent sexual and reproductive health services in sub-saharan africa: a qualitative systematic review. Open Access Journal of Contraception. 2023:103–18.

23. Sanyang Y, Sanyang S, Ladur AN, Cham M, Desmond N, Mgawadere F. Are facility service delivery models meeting the sexual and reproductive health needs of adolescents in Sub-Saharan Africa? A qualitative evidence synthesis. BMC health services research. 2025;25(1):193.

24. Hensen B, Floyd S, Phiri MM, Schaap A, Sigande L, Simuyaba M, et al. The impact of community-based, peer-led sexual and reproductive health services on knowledge of HIV status among adolescents and young people aged 15 to 24 in Lusaka, Zambia: The Yathu Yathu cluster-randomised trial. PLoS medicine. 2023;20(4):e1004203.

25. Shah P, Kibel M, Ayuku D, Lobun R, Ayieko J, Keter A, et al. A pilot study of “peer navigators” to promote uptake of HIV testing, care and treatment among street-connected children and youth in Eldoret, Kenya. AIDS and Behavior. 2019;23(4):908–19.

26. Borji-Navan S, Maleki N, Keramat A. Efficacy of digital health interventions used for adolescent’s sexual health: An umbrella review. Health science reports. 2024;7(12):e70212.

27. Belay HG, Arage G, Degu A, Getnet B, Necho W, Dagnew E, et al. Youth-friendly sexual and reproductive health services utilization and its determinants in Ethiopia: A systematic review and meta-analysis. Heliyon. 2021;7(12).

28. Di Iorio R. Exploring the Feasibility and Acceptability of a Participatory Workshop Program to Reduce Stigma Associated with Sexual and Reproductive Health and Contraception Among Adolescent Girls and Young Women in Kilimanjaro, Tanzania: A Mixed-Methods Pilot Study: Queen’s University (Canada); 2025.

29. Cutherell M, Phillips M, Ellett C, Woubishet E, Ede JO, Adesina A, et al. Balancing evidence-informed and user-responsive design: Experience with human-centered design to generate layered economic empowerment and SRH programming in Tanzania, Ethiopia, and Nigeria. Gates Open Research. 2023;7:106.

30. Njoki N, Cutherell M, Musau A, Mireri D, Nana-Sinkam A, Phillips M. Applying human-centered design to replicate an adolescent sexual and reproductive health intervention: a case study of Binti Shupavu in Kenya. Global Health: Science and Practice. 2023;11(6).

31. Svanemyr J, Amin A, Robles OJ, Greene ME. Creating an enabling environment for adolescent sexual and reproductive health: a framework and promising approaches. Journal of adolescent health. 2015;56(1):S7–S14.

32. Krug C, Neuman M, Rosen JE, Weinberger M, Wallach S, Lagaay M, et al. Effect and cost-effectiveness of human-centred design-based approaches to increase adolescent uptake of modern contraceptives in Nigeria, Ethiopia and Tanzania: population-based, quasi-experimental studies. PLOS Global Public Health. 2023;3(10):e0002347.

33. Maryam S, Nonhlanhla O, Natsayi C, Thembelihle Z, Sakhile M, Nondumiso M, et al. Thetha Nami: participatory development of a peer-navigator intervention to deliver biosocial HIV prevention for adolescents and youth in rural South Africa. 2021.

