## Supplemental Data 1 for "Co-Designing an Integrated Intervention Package to Strengthen Adolescent Sexual and Reproductive Health Services in Primary Healthcare Units in Southern Ethiopia: A Human-Centered Design Approach"

### Appendixes

**Table 1:** Integrated Priority Barriers to Adolescent SRH Service Utilization

| Thematic Group | Identified Barriers | Description |
|---|---|---|
| **1. Service Delivery** | 1. Lack of adolescent-friendly and confidential services<br>2. Frequent stockouts of essential SRH commodities (e.g., condoms, STI kits, emergency contraceptives)<br>3. Inadequate provider training and judgmental attitudes<br>4. Long waiting times and inconvenient service hours<br>5. Weak referral and follow-up mechanisms<br>6. Services denied based on age or parental consent<br>7. Use of multi-purpose rooms limiting privacy<br>8. Absence of youth-focused outreach or mobile services | Services are not tailored to adolescent needs, with poor confidentiality and access interruptions; provider skills and attitudes limit quality; access barriers exist due to timing, policies, and infrastructure; outreach is insufficient. |
| **2. Family and Community Engagement** | 1. Lack of parent-adolescent communication on SRH topics<br>2. Unsupportive or judgmental family environments<br>3. Community stigma linking SRH service use to promiscuity<br>4. Cultural taboos and harmful norms around adolescent sexuality<br>5. Low community acceptability of SRH services for unmarried adolescents<br>6. Misinformation or religious opposition to SRH topics | Poor family dialogue and unsupportive environments reduce adolescent knowledge and service use; community stigma and taboos limit open discussion and acceptance; misinformation undermines education and uptake. |
| **3. Information and Education** | 1. Lack of basic knowledge on SRH and available services<br>2. SRH topics skipped or poorly addressed in school curricula<br>3. Adolescents unaware of their rights to access care<br>4. Information not age-appropriate, culturally relevant, or youth-friendly<br>5. Misinformation spread through peers and social media without correction | Adolescents lack foundational SRH awareness and rights knowledge; education delivery is inadequate or mismatched; misinformation contributes to confusion and myths. |

**Table 2: Priority Adolescent Sexual and Reproductive Health Problems Identified by Thematic Area**

| Thematic Area | Priority Problem |
|---|---|
| **Service Delivery** | 1. **Problem 1:** Lack of adolescent-friendly services |
| | 2. **Problem 2:** Absence of adolescent-focused outreach services |
| | 3. **Problem 3:** Long waiting times and inconvenient service hours |
| | 4. **Problem 4:** Weak monitoring, evaluation, and accountability mechanisms |
| **Family & Community Engagement** | 5. **Problem 1:** Poor parent–adolescent communication on SRH issues |
| | 6. **Problem 2:** Low community acceptance of adolescent SRH needs |
| | 7. **Problem 3:** Misinformation and religious opposition to adolescent SRH |
| **Information & Education** | 8. **Problem 1:** Lack of basic knowledge of SRH services among adolescents |
| | 9. **Problem 2:** SRH topics are inadequately addressed in the school curriculum |
| | 10. **Problem 3:** Widespread misinformation spread through social media platforms |

**Table 3:** Summary of Ideas Generated by Participants and Their Thematic Clustering

| Thematic Category | Number of Ideas | Key Focus Areas |
|---|---|---|
| Service Delivery | 71 | Enhancing access, youth-friendliness, provider capacity, and service quality |
| Family & Community Engagement | 40 | Strengthening parental involvement, community support, and cultural responsiveness |
| Information & Education | 29 | Improving knowledge, awareness, and communication channels |

**Table 4:** Feasibility and Impact Assessment of ASRH Intervention Themes

| Intervention Theme | Feasibility | Impact | Key Strengths | Challenges | Implementation Priority |
|---|---|---|---|---|---|
| Service Delivery Strengthening | High | High | Actionable, infrastructure-ready, scalable | Training & supervision; resource allocation | Phase 1 (Top Priority) |
| IEC Strategies | Moderate-High | Moderate-High | Scalable; adaptable to digital/peer-led platforms | Digital access disparities; literacy gaps | Phase 1–2 |
| Family & Community Engagement | Moderate | High | Fosters supportive norms; community ownership | Requires long-term behavioral change | Phase 2 onward |

**Table 5: Final Co-Designed Adolescent Sexual and Reproductive Health Intervention Packages Designed in South Ethiopia, 2025.**

| Intervention Package | Target Population | Key Barrier Addressed | Core Components / Activities | Delivery Agents | Setting | Frequency / Dose | Co-Design Contribution |
|---|---|---|---|---|---|---|---|
| **Capacity-Building for Providers and Managers** | Health providers & facility managers | Limited provider skills, judgmental attitudes, poor client-centered care | Workshops, mentorship, refresher sessions, supervision & feedback | Trainers and senior clinicians. | Health facilities | Initial 5-day workshop + ongoing mentorship | Stakeholders emphasized non-judgmental, client-centered care and practical, hands-on training |
| **Adolescent-Friendly Service Environments** | Adolescents (10–24 yrs) | Unwelcoming facilities, limited privacy, inconvenient hours | Facility redesign, extended hours, adolescent-specific service blocks, educational materials | Facility managers, health care providers | Health facilities | Continuous | Adolescents highlighted privacy, comfort, and visual cues for inclusivity |
| **Integrated Peer Navigation** | Adolescents (10–24 yrs) | Low awareness & engagement with SRH services | Peer counseling, community outreach, facility support, referral pathways | Trained peer navigators | Community & facility | Initial 3-day workshop +Ongoing, structured follow-ups | Adolescents suggested peer-led engagement and follow-up for continuous support |
| **SRH Call Center Support** | Adolescents (10–24 yrs) | Lack of timely information, stigma, distance barriers | Toll-free hotline, counseling, information dissemination, appointment scheduling, referral | Call center staff, integrated with facilities | Remote (phone-based) | As needed | Adolescents requested confidential, anonymous, and convenient access to SRH information |
